# Decreases in liver cT1 accurately reflect histological improvement induced by therapies in MASH: a multi-centre pooled cohort analysis

**DOI:** 10.1101/2024.02.28.24302571

**Authors:** Naim Alkhouri, Cayden Beyer, Elizabeth Shumbayawonda, Anneli Andersson, Kitty Yale, Timothy Rolph, Raymund Chung, Raj K Vuppalanchi, Kenneth Cusi, Rohit Loomba, Andrea Dennis, Michele Pansini

## Abstract

**Background & Aims:** Iron corrected T1 (cT1) is an MRI derived biomarker of liver disease activity. Emerging data suggest a change in cT1 of ≥ 80 ms reflects histological improvement. We aimed to validate the association between the ≥ 80 ms decline in cT1 and histological improvement, specifically the resolution of MASH.

**Methods:** A retrospective analysis of study participants from three interventional clinical trials with histologically confirmed MASH (n = 150) who underwent multi-parametric MRI to measure cT1 (LiverMultiScan®) and biopsies at baseline and end of study. Histological responders were defined using the four criteria: (1) a decrease in NAFLD Activity score (NAS) ≥ 2 with no worsening in fibrosis, (2) a decrease in fibrosis ≥ 1 stage with no worsening in NAS, (3) both a NAS decrease ≥ 2 and a fibrosis decrease ≥ 1, and (4) MASH resolution with no worsening in fibrosis. Difference in the magnitude of change in cT1 between responders and non-responders was assessed.

**Results:** Significant decreases in cT1 were observed in responders for all the histological criteria. The largest decrease was observed for those achieving MASH resolution, and was 119ms, compared to 43ms for non-responders. The optimal reduction in cT1 for separating responders from non-responders for MASH resolution was −74ms (64ms-73ms for the other criteria), in close agreement with the previously predefined threshold of −80ms. Those achieving an ≥ 80 ms reduction in cT1 were substantially more likely to achieve histological response with odds ratios ranging from 2.7 to 6.3.

**Conclusions:** These results demonstrate that a reduction in cT1 of 80 ms was associated with histological response supporting the utility of cT1 to predict clinical improvement in patients undergoing therapeutic intervention.

## INTRODUCTION

Metabolic Dysfunction-Associated Steatotic liver disease (MASLD) affects 30% of the population and is a growing concern worldwide^1^ with up to 14% of the middle-aged population^2^ progressing to a more severe condition, metabolic associated steatohepatitis (MASH). MASH is characterized by the presence of liver steatosis, inflammation and ballooning, in conjunction with one cardiometabolic risk factor, and is becoming the leading cause for liver transplantation^3, 4^ and hepatic cancer.^5^ There is currently no approved therapy for MASH; however, there are several ongoing interventional trials of pharmacotherapies targeting various pathogenic processes underlying the disease.^6–12^ Currently available interventions which have demonstrated positive effects in patients with MASH involve lifestyle changes with diet and exercise, vitamin E and treatments for comorbidities such as diabetes and obesity.^13, 14^

Monitoring of patients and assessing the efficacy of interventions is critical to reducing the prevalence of MASH. Non-invasive tests that provide precise measurement and the ability to dynamically detect early changes in the disease-driving features is of prime interest in the management of patients, as well as for efficacy assessment in clinical trials. However, regulatory agencies currently require liver biopsy in phase 3 MASH trials as the primary endpoint for efficacy assessment. Liver biopsy is unsuitable and impractical as a reference standard for longitudinal monitoring^15^ in clinical trials. It is an invasive procedure yielding highly variable results and is associated with clinical complications and significant costs^15–17^ making it impractical for widespread clinical practice. Efforts to identify alternative, non-invasive robust biomarkers for detecting change following treatment, and as surrogate endpoints, have been strongly encouraged by regulatory bodies such as the United States Food and Drug Administration (FDA)^18^.

Multiparametric magnetic resonance imaging (mpMRI) is a safe alternative to liver biopsy for tissue characterisation in clinical trials. Unlike blood tests, imaging methods are inherently liver specific, and unlike biopsies, they quantify the health status of the entire liver. MRI proton density fat fraction (PDFF) is a reliable and accurate measure of liver fat and several studies have examined the association of change in PDFF with histological response ^19–22^. A relative decline in PDFF of ≥ 30% has been associated with an increased probability of meeting the histological endpoints of a 2-point change in the NAFLD activity score (NAS) with no worsening in fibrosis (with odds ratio of (OR) 4.86) and resolution of MASH with no worsening in fibrosis (OR 2.2) ^21^. Regarding improvement in fibrosis however, PDFF is challenging to interpret. PDFF has a non-linear association with fibrosis. There is an initial increase in PDFF as fibrosis develops to F2, then a gradual decline.^23–25^ MASH cirrhosis is characterised by progressive reduction in liver fat content as the fibrosis burden increases and consequently the development of cryptogenic cirrhosis. This is likely the main reason that liver fat content has not been shown to predict liver-related outcomes^26, 27^.

MRI may offer another parameter, Iron-corrected T1 mapping (cT1), which could be used as either an alternative or to complement PDFF. cT1 is derived from a signal from the T1-mapping approach^28^ which has been corrected for MRI field strength and the presence of liver iron.^29^ It is a measure of water content in the tissue, and thus is sensitive to both the presence of inflammation and increases in extracellular collagen matrix. cT1 has been used in the diagnosis and monitoring of MASLD and MASH patients who are at high-risk of progression to adverse clinical outcomes.^30^ It has been shown to correlate with histologic features of the NAS as well as fibrosis grading.^29, 31, 32^ Elevated values of cT1 has been shown to be predictive of both major liver^33^ and cardiac^34^ related clinical outcomes.

Several trials are already employing cT1 as a diagnostic screening biomarker as well as a secondary or exploratory endpoint in interventional trials for patients with MASH.^35–37^ Decreases in cT1 have been observed after bariatric surgery^38^, low energy diets^13^, and following treatment with investigational drugs targeting liver specific fat reduction^39, 40^ and fibrosis reduction^41, 42^, with changes observed as early as 12-weeks.^41^ A previous cross-sectional investigation estimated that a decrease in cT1 of ∼80 ms corresponded to a 2-point decrease in the NAS with no worsening in fibrosis^43^, indicative of a clinically significant improvement.

In this analysis of data pooled from three interventional phase 2 MASH trials, we aimed to (1) validate previous observations and establish the magnitude of change in cT1 that correspond to histological endpoints indicative of positive improvement in liver health, in particular MASH resolution, and (2) determine whether an 80 ms drop in cT1 predicts the likelihood of reaching a histological response.

## PATIENTS AND METHODS

### Design and Study Participants

This study was a retrospective, pooled longitudinal analysis of three independent interventional MASH clinical trials run between 2015–2021. All three studies were phase 2, multi-centre, MASH clinical trials in adult US populations recruiting from secondary care and included different treatment doses as well as placebo treated participants. All the clinical investigations were conducted in accordance with the Declaration of Helsinki 2013, approved by local relevant institutional review boards and written informed consent was obtained from all participants. Study 1 (n = 18), scanned and biopsied enrolled patients at baseline and 12 weeks post-treatment. Inclusion to the study (NCT02443116) required definite histological evidence of MASH (NAS ≥ 4 with a score ≥ 1 for each component [steatosis, lobular inflammation, and ballooning], and biopsy-scored fibrosis stage 1-3) and a PDFF ≥ 8%. Exclusion criteria included previous liver transplant or an inability to undergo MRI or biopsy. For Study 2 (n = 42), enrolled patients were scanned and biopsied at baseline and 22–24 weeks post-baseline, after 16 weeks treatment. Only patients who achieved a ≥ 30% relative reduction in PDFF after 12 weeks treatment were biopsied at 22-24 weeks. Inclusion to the study (NCT03976401) required definite histological evidence of MASH and a PDFF ≥ 10%, while exclusion criteria included significant alcohol consumption, an inability to undergo MRI or biopsy, or planning to undergo or having undergone liver transplant or bariatric surgery. Study 3 (n = 140), enrolled patients were scanned and biopsied at baseline and after 52 weeks treatment. Inclusion to the study (NCT03551522) required definite histological evidence of MASH and a PDFF ≥ 10%, while exclusion included significant alcohol consumption, inability to undergo MRI or biopsy, or a BMI < 18.5 kg/m^2^.

### Histological assessment

Histology was graded according to the NASH Clinical Research Network (NASH-CRN) for Kleiner-Brunt fibrosis, hepatocellular ballooning, lobular inflammation, steatosis, and the composite NAS. All pathologists were blinded to all clinical data including patient characteristics and non-invasive assessment data. Biopsy scores used for the analysis were those collected and centrally read as part of each independent study, further pooled central re-reading was not performed.

### Multiparametric Imaging

All participants underwent abdominal multiparametric MRI examination with the LiverMultiScan image acquisition protocol, which was installed, calibrated, and both phantom and volunteer tested on all the MRI systems according to manufactures guidance.^44^ LiverMultiScan reports cT1, PDFF and T2* as a marker of liver iron. Patients underwent MRI having fasted for at least 4 hours. The average scan time for this protocol was 10 minutes. The MRI protocol has been described elsewhere.^44^ To enable a median value across the liver to be calculated, generated cT1 maps were delineated into whole liver segmentation maps using a semi-automatic method, with non-parenchyma structures such as bile ducts and large blood vessels as well as image artifacts excluded. All image analysis was completed by analysts blinded to the clinical data.

### Statistical Analysis

All statistical analyses were performed in R Studio version 4.2.2. with a p-value of < 0.05 considered significant. Mean and standard deviation (SD) were used to describe all normally distributed continuous variables. Median and interquartile range (IQR) were used to describe all non-normally distributed continuous variables. Frequency and percentage were used to describe categorical variables. Case wide omission of patients missing histology scores (NAS or fibrosis) or cT1 values from either timepoint (baseline or follow-up) was performed.

Patients were characterized as responders or non-responders based on the histological changes reported via pathology. Responders were classified under four different criteria: (1) a NAS decrease ≥ 2 with no worsening in fibrosis, (2) a fibrosis decrease ≥ 1 with no worsening in NAS, (3) both a NAS decrease ≥ 2 and a fibrosis decrease ≥ 1, and (4) MASH resolution - a ballooning score of 0 and inflammation of ≤ 1 at follow-up, with no worsening in fibrosis. Non-responders were classified as patients who did not respond under any of the four criteria mentioned above, unless otherwise stated.

Receiver operating characteristic curves were used to evaluate the diagnostic accuracy of change in cT1 to classify participants as either histological responder or non-responder, for each of the four classifications. Youden’s index was used to estimate the optimal threshold and median changes in responders reported to illustrate the observed difference between responders and non-responders. To validate the previously estimated meaningful change on 80ms for identifying responders, sensitivity and specificity was calculated, and odds ratio was used to define the probability of responding on histology. Average change in cT1 between responders and non-responders was assessed using Wilcoxon rank sum test.

## RESULTS

### Cohort description

In the pooled independent participant data set used for this study, following omission of cases missing histology or cT1 data, a cohort of n = 150 was included for analysis (**Figure 1**, **Table 1**). The descriptive statistics in Table 1 (and **Supplementary Table 1,2**) show that the histology (steatosis, inflammation, ballooning, and fibrosis) and NAS scores were comparable between the three individual studies. Figure 2 shows representative quantitative cT1 maps of the liver in histological responders and non-responders.

**Figure 1:**
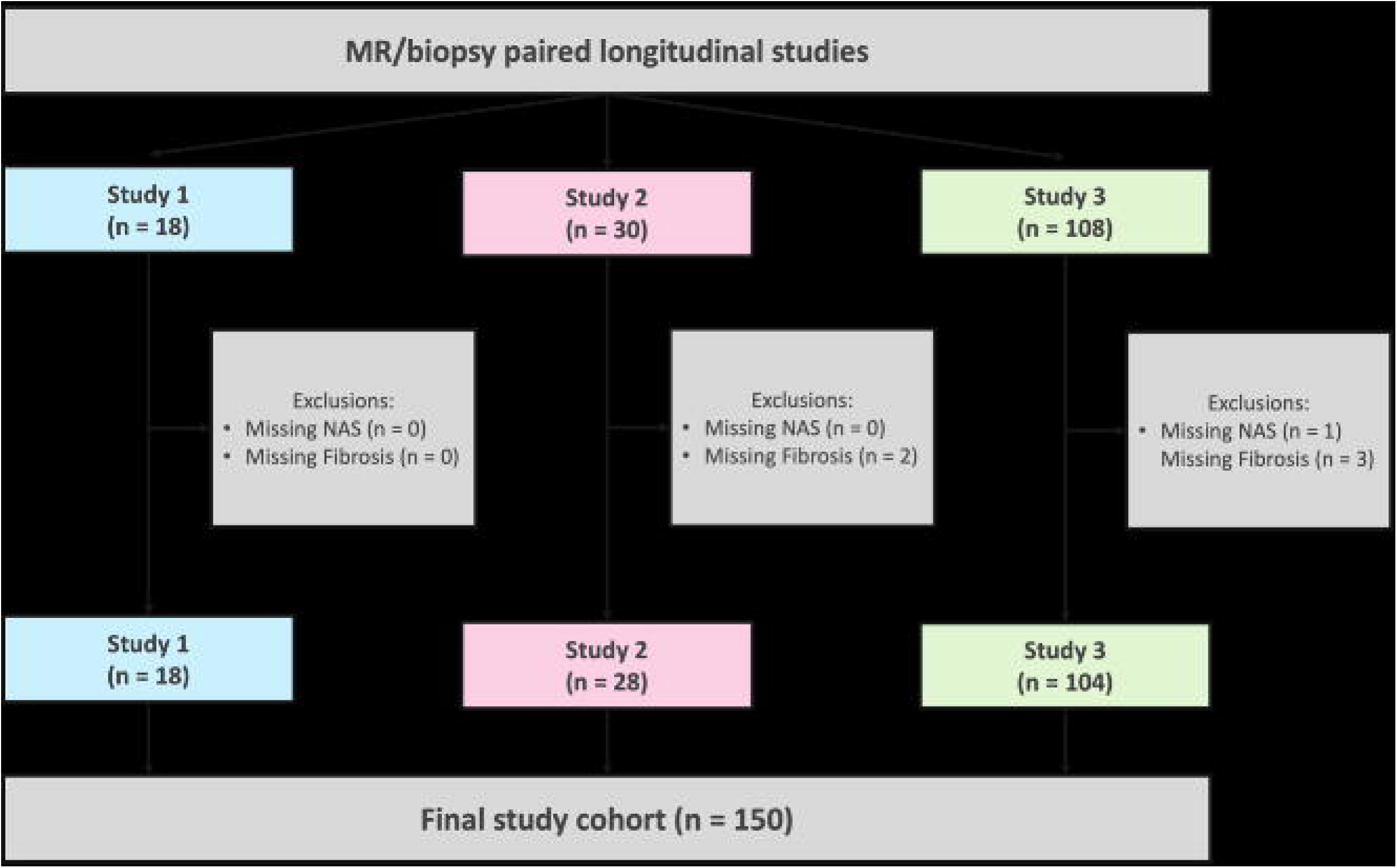
Consort diagram of subject omissions due to missing data for each of the three pooled studies.

**Figure 2:**
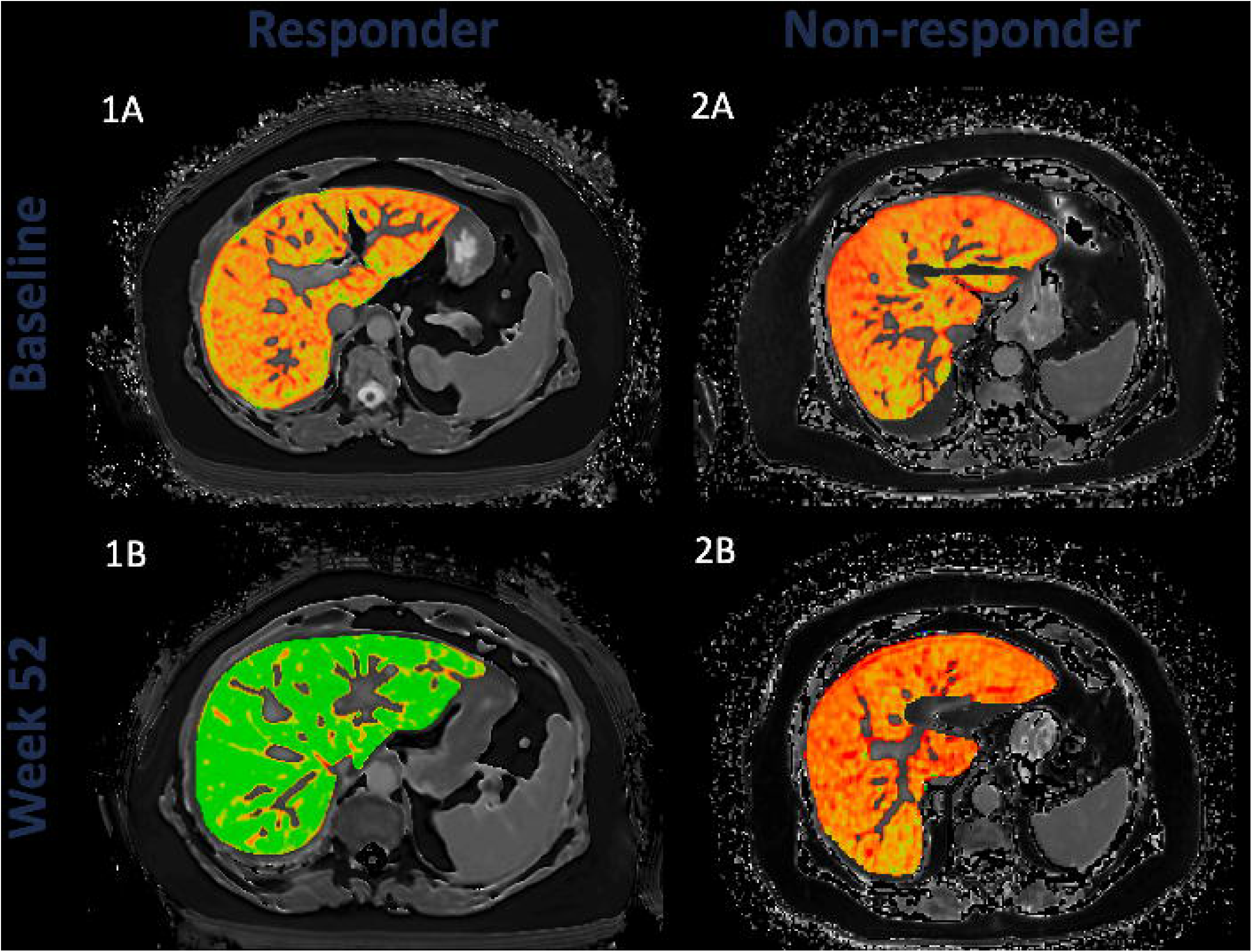
Example images of cT1 and PDFF maps for comparison. cT1 maps are shown at baseline (A) and week 52 follow-up (B). PDFF maps at baseline (C) and 52-week follow-up (D). The images depicted for three patients who were characterized as a cT1 responder, PDFF responder and histological responder (1); cT1 non-responder, PDFF non-responder, and histological non-responder (2); and cT1 non-responder, PDFF responder, and histological non-responder (3). cT1 responders and non-responders classified as having a cT1 decrease ≥ 80 ms and a cT1 decrease < 80 ms, respectively. PDFF responders and non-responders classified as having a PDFF decrease ≥ 30% and a PDFF decrease < 30%, respectively. Histological responders defined as reaching MASH resolution.

**Table 1:** Patient demographics and MRI/histology baseline characteristics in the whole cohort and sub-grouped by cT1 change from baseline value. Continuous variables compared across subgroups using Wilcoxon rank sum test; normally distributed variables described as Mean (SD), non-normally distributed described as Median (IQR). Categorical variables described as Frequency (%); variables containing any entry with a value < 5 were compared using Fisher’s Exact test, variables with all entries ≥ 5 were compared using the Chi-square test.

| Characteristic | All patients (n=150) | cT1 responders (cT1 change ≥80ms), n= 53 | cT1 non-responders (cT1 change <80ms), n= 97 | P value; responder vs. non-responder |
| --- | --- | --- | --- | --- |
| Age (years) | 57 (17) | 57 (17) | 53 (18) | 0.47 |
| Sex (% Female) | 102 (68%) | 69 (71%) | 33 (62%) | 0.27 |
| BMI (kg/m<sup>2</sup>) | 36.0 (9.8) | 35.0 (10.0) | 37.4 (10.3) | 0.19 |
| Ethnicity |  |  |  | 0.18 |
| Hispanic/Latino | 62 (41%) | 44 (45%) | 18 (34%) |  |
| Non-Hispanic/Latino | 88 (59%) | 53 (55%) | 35 (66%) |  |
| Diabetes | 56 (37%) | 33 (34%) | 23 (43%) | 0.26 |
| cT1 (ms) | 919 (133) | 890 (141) | 955 (156) | **0.001** |
| PDFF (%) | 18.8 (7.6) | 17.5 (6.1) | 20.8 (6.7) | **0.003** |
| Steatosis |  |  |  | **0.014** |
| 0 | 0 (0%) | 0 (0%) | 0 (0%) |  |
| 1 | 48 (32%) | 39 (40%) | 9 (17%) |  |
| 2 | 66 (44%) | 37 (38%) | 29 (55%) |  |
| 3 | 36 (24%) | 21 (22%) | 15 (28%) |  |
| Inflammation |  |  |  | **0.017** |
| 0 | 10 (6.7%) | 7 (7.2%) | 3 (5.7%) |  |
| 1 | 81 (54%) | 60 (62%) | 21 (40%) |  |
| 2 | 52 (35%) | 28 (29%) | 24 (45%) |  |
| 3 | 7 (4.7%) | 2 (2.1%) | 5 (9.4%) |  |
| Ballooning |  |  |  | 0.68 |
| 0 | 4 (2.7%) | 2 (2.1%) | 2 (3.8%) |  |
| 1 | 73 (49%) | 46 (47%) | 27 (51%) |  |
| 2 | 73 (49%) | 49 (51%) | 24 (45%) |  |
| Fibrosis |  |  |  | 0.49 |
| 0 | 0 (0%) | 0 (0%) | 0 (0%) |  |
| 1 | 28 (19%) | 17 (18%) | 11 (21%) |  |
| 2 | 50 (33%) | 34 (35%) | 16 (30%) |  |
| 3 | 64 (43%) | 39 (40%) | 25 (47%) |  |
| 4 | 8 (5.3%) | 7 (7.2%) | 1 (1.9%) |  |
| NAS |  |  |  | 0.1 |
| 0 | 0 (0%) | 0 (0%) | 0 (0%) |  |
| 1 | 0 (0%) | 0 (0%) | 0 (0%) |  |
| 2 | 4 (2.7%) | 4 (4.1%) | 0 (0%) |  |
| 3 | 20 (13%) | 17 (18%) | 3 (5.7%) |  |
| 4 | 36 (24%) | 24 (25%) | 12 (23%) |  |
| 5 | 52 (35%) | 33 (34%) | 19 (36%) |  |
| 6 | 28 (19%) | 13 (13%) | 15 (28%) |  |
| 7 | 7 (4.7%) | 4 (4.1%) | 3 (5.7%) |  |
| 8 | 3 (2.0%) | 2 (2.1%) | 1 (1.9%) |  |

### cT1 change associated with clinically meaningful histological response

The drop in cT1 in patients who met any of the 4 criteria was significantly different from those who did not respond (figure 3). The median (IQR) decreases in cT1 in the patients who met the histological response criteria of MASH resolution was 119 ms (± 45 ms), versus 43 ms (± 50 ms) for those who failed to meet any histological response criteria (**Figure 3**). The Youden’s index for this criterion was −74ms (sensitivity 0.73, specificity 0.75). Evaluation of each of the other three individual histological response criteria showed that improvement in cT1 was consistently associated with positive histological response (**Figure 3**).

**Figure 3:**
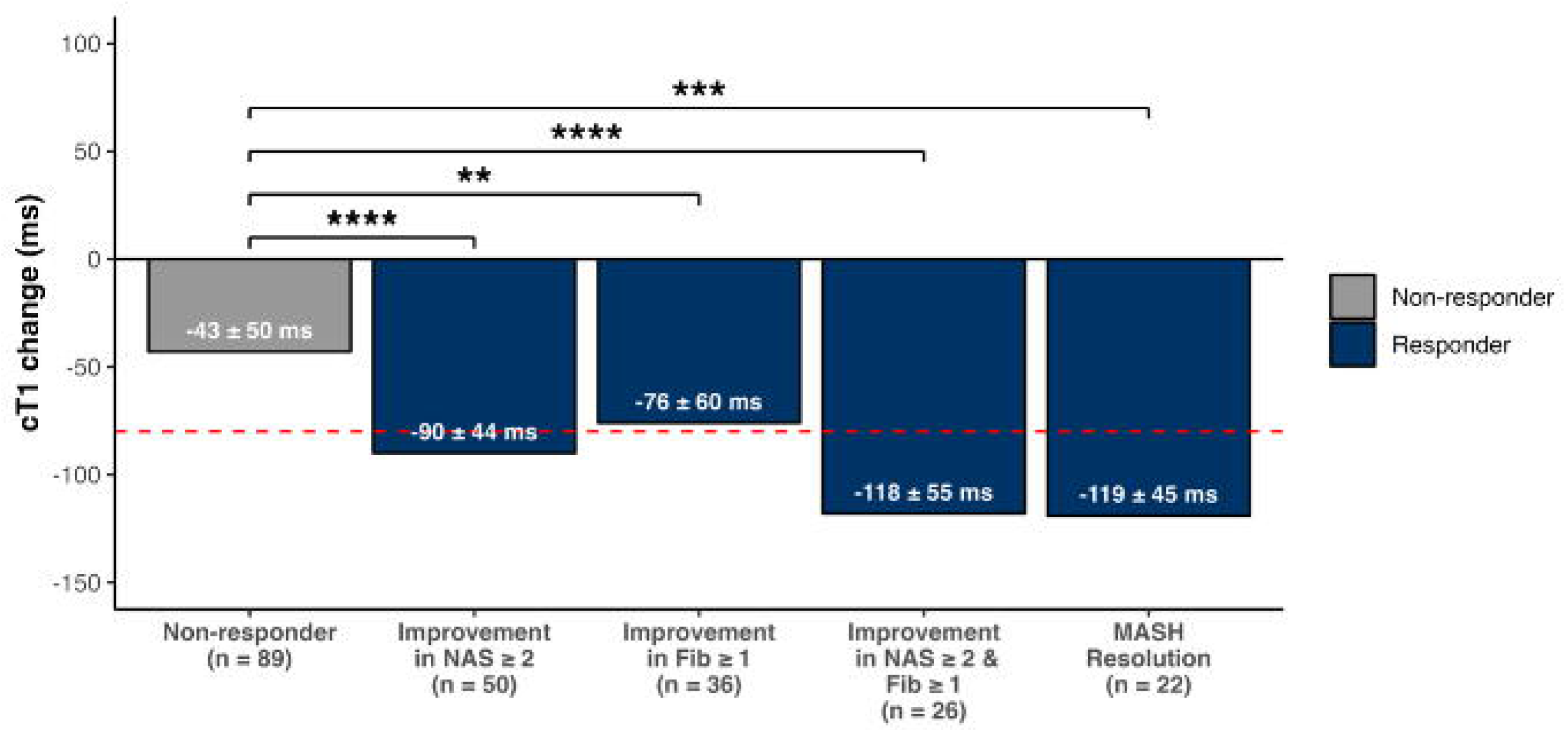
Median cT1 changes in responders, and non-responders. Median cT1 values are shown for each of the four different criteria for histological response, and the corresponding p-value (Wilcoxon rank sum test).

### Using cT1 to identify positive histological responders

Change in cT1 had good diagnostic accuracy to identify patients with MASH resolution (AUC = 0.76 [95% CI: 0.63–0.88]), with performance consistent across all response criteria. To measure response, a threshold of cT1 ≥ 80 ms had a sensitivity of 0.68 (ranging between 0.47–0.68 across individual criteria), and an average specificity of 0.75, with positive predictive values ranging from 0.41–0.58 and negative predictive values from 0.77–0.91 (**Table 2**).

**Table 2:** Diagnostic accuracy of cT1 threshold of 80 ms and for the Youden’s index for identifying histological responders under each of the response criteria. Diagnostic accuracy metrics include sensitivity, specificity, positive predictive value (PPV), and negative predictive value (NPV).

| Responder Definition | Threshold | Sensitivity | Specificity | PPV | NPV |
| --- | --- | --- | --- | --- | --- |
| Improvement NAS $\geq 2$ & no worsening fibrosis | 80ms | 0.60 | 0.75 | 0.58 | 0.77 |
|  | 64ms | 0.74 | 0.69 | 0.57 | 0.82 |
| Improvement fibrosis $\geq 1$ & no worsening in NAS | 80ms | 0.47 | 0.75 | 0.44 | 0.78 |
|  | 73ms | 0.56 | 0.75 | 0.48 | 0.81 |
| Improvement NAS $\geq 2$ & Fibrosis $\geq 1$ | 80ms | 0.62 | 0.75 | 0.42 | 0.87 |
|  | 68ms | 0.77 | 0.72 | 0.44 | 0.91 |
| NASH resolution & no worsening in Fibrosis | 80ms | 0.68 | 0.75 | 0.41 | 0.91 |
|  | 74ms | 0.73 | 0.75 | 0.42 | 0.92 |

Those participants with a cT1 decrease of ≥ 80 ms following intervention had a higher odds ratio of meeting the histological response criteria, particularly those having MASH resolution with no worsening in fibrosis (**Figure 4**).

**Figure 4:**
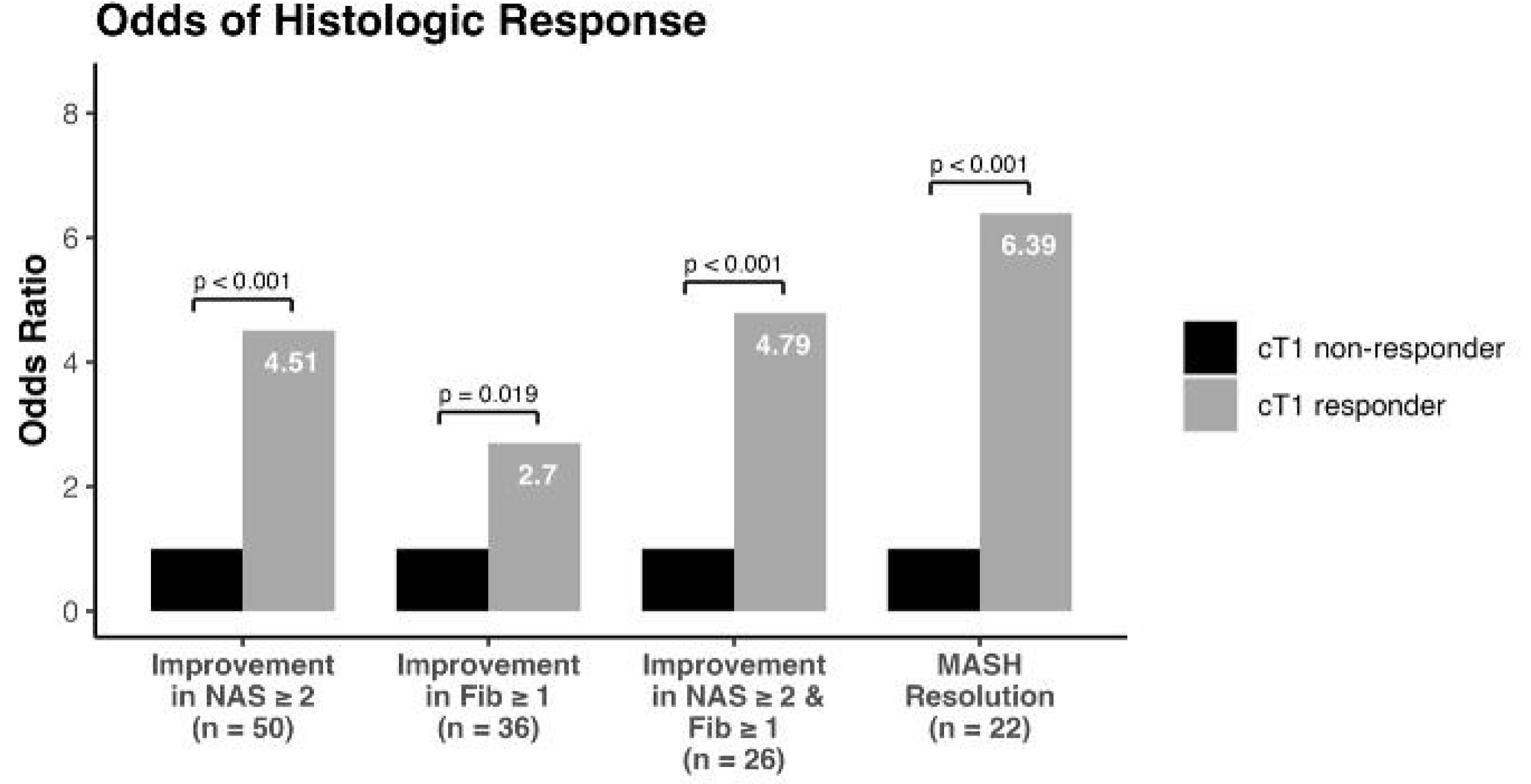
Odds ratios and corresponding p-values (Fisher’s exact test) for the percentage of patients with cT1 decrease ≥ 80ms meeting histologic response criteria (responders) under each of the histological response criteria

## DISCUSSION

The aim of this analysis was to confirm the magnitude of change in cT1 at an individual patient level that corresponds with surrogate histological endpoints used in MASH trials. Following previous cross-sectional estimates and observations for other studies^32, 36^ a decrease in cT1 of ∼80 ms appeared reasonably likely to predict histological improvement.

In this work, four definitions of histological response were used to validate the clinical relevance of a cT1 threshold: NAS decrease ≥ 2 with no worsening in fibrosis, a decrease in fibrosis ≥ 1 with no worsening in NAS, NAS decrease ≥ 2 with decrease in fibrosis ≥ 1 and MASH resolution with no worsening of fibrosis. The Youden’s index for these criteria were 64ms, 73ms, 68ms and 74ms respectively, in close agreement with a predefined threshold of 80ms. Our findings showed that a patient with a reduction in cT1 ≥ 80 ms was over six times more likely to have MASH resolution than patients with a cT1 change below this threshold. Thus, for the purpose of drug development an average change in cT1 could effectively be used to assess efficacy and likelihood of a response in terms of the histological endpoints. For clinical application on an individual level, a change in cT1 of 80 ms or greater had negative predictive values of between 0.77–0.91, suggesting good performance to rule out those patients who are not responding to treatment and who may benefit from stopping.

Currently FDA requires liver histology as surrogate endpoints for approval of a medicine for treating MASH. The well reported limitations of biopsy (variability in sample location, inter-rater variability, risk of major complications of 2.4%^15^) suggest the reliance for confirmation of disease in clinical trials for regulatory submission would be impractical in everyday clinical practice. Additionally, the slow progression of MASH and the relatively high frequency of disease regression, preclude liver-related clinical outcomes as practical endpoints for interventional trials in MASH, except for late-stage cirrhosis cohorts.^45^ As a result, regulatory bodies, clinicians, drug developers and patient advocacy groups are seeking non-invasive alternatives.^46^ As yet no blood biomarkers and associated scoring systems have met desired thresholds for sensitivity and specificity.^47^ Whilst cT1 is not specific to individual features of histology, it does detect improvements in liver health, which may be of more importance than the mechanism of change in clinical practice. When considering a target magnitude change in cT1, healthy liver has a cT1 value of 800 ms or less.^30^ A reduction of 80 ms or more will help as an effective interpretation marker to track how a patient is responding; however, the goal of treatment should ultimately be to achieve a cT1 of 800 ms. Several studies are incorporating an MRI sub study as an orthogonal, non-invasive measure to corroborate histology results. This data may help inform how imaging in phase 3 trials could be used to determine who should have a confirmatory liver biopsy to assess response, reducing the need for everyone to undergo the invasive procedure. For example, it might be reasonable to suggest that only those who did not reach an 80 ms drop would be suitable for biopsy, e.g. 65% of enrolled patients from this cohort.

Despite the absence of any pharmacological interventions, a significant proportion of patients in the placebo arm appear to show improvements in liver histology^48^ and based on imaging measurements. This has been attributed to the possibility that MASH can regress spontaneously, but more likely reflects the Hawthorne effect among participants in interventional trials.^49^ Reflecting the noise inherent to measuring cT1, a change in cT1 of ± 46 ms has previously been determined to be the smallest detectable difference based on test-retest of healthy participants repeatedly scanned within the same session.^44^ A larger change may be observed under less controlled conditions or across different days because of more physiological noise. From a practical perspective, to adequately power an imaging sub-study based on cT1 to detect a statistical difference between patients on treatment compared to placebo, a desired effect size (difference between groups) of 40-50 ms may be advised.

A strength of this retrospective analysis was the use of three independent international clinical trials evaluating the efficacy of different MASH pharmacotherapies using standardised imaging protocols. By rigorously investigating the performance of MRI to distinguish histological responders from non-responders, based on a range of response criteria, these results provide a robust and comprehensive assessment of the ability of cT1 to identify clinically significant changes in a patient’s liver health. These findings also show the generalisability of the change in cT1 following intervention supporting a change of ≥ 80 ms as being clinically meaningful.

While histology remains the only currently accepted endpoint for liver health assessment in MASH, we did not execute another central read and thus assumed that the expertise of pathologists conducting the central reads across all included clinical trials was equivalent. We acknowledge the limitation this adds to this investigation. However, as the field moves towards non-invasive measures as potential surrogate biomarkers of response, the primary aim of this investigation was the validation of cT1 thresholds. Future studies should investigate the relationship between non-invasive biomarkers (including blood markers) which can be used in combination to comprehensively define changes in overall health following treatment. Insights gained from such combinations could enable understanding of the impact of improving liver health on whole body metabolic features such as body composition, weight, and fat free mass. This will be even more relevant as multiple treatment options emerge for patients with MASLD and concomitant diabetes allowing to better-selection of the most appropriate first line treatment and/or informing on stopping rules.

In summary, this study substantiates that a decrease in cT1 by 80 ms or more serves as a reliable indicator with tangible histological benefits in MASLD patients. Amid the increasing focus on precision medicine and the imperative for dependable, standardized measures, our findings underscore the value of non-invasive mpMRI markers in guiding patient care. Furthermore, these markers hold promise for shaping public health initiatives aimed at alleviating the burden of MASLD in the foreseeable future. This becomes especially vital in clinical environments where timely and accurate monitoring can profoundly influence both the efficacy of treatment and the patient’s overall well-being.

## Clinical trial number(s)

NCT02443116, NCT03976401, NCT03551522

## Data Availability

All data produced in the present work are contained in the manuscript

## ABBREVIATIONS

AUC: area under the curve
AUROC: area under the operator characteristic curve
cT1: iron-corrected T1-mapping
FDA: food and drug administration
IQR: interquartile range
MASH: metabolic dysfunction-associated steatohepatitis
MASLD: metabolic dysfunction-associated steatotic liver disease
mpMRI: multi-parametric magnetic resonance imaging
NAS: NAFLD activity score
NASH-CRN: NASH clinical research network
OR: odds ratio
PDFF: proton density fat-fraction
ROC: receiver operator characteristic curve.

## ACKNOWLEDGEMENTS

The investigators thank the patients for trial participation.

## REFERENCES

1. Cotter TG, Rinella M. Nonalcoholic Fatty Liver Disease 2020: The State of the Disease. Gastroenterology 2020;158:1851–1864. doi: 10.1053/j.gastro.2020.01.052

2. Harrison SA, Gawrieh S, Roberts K, Lisanti CJ, Schwope RB, Cebe KM, et al. Prospective evaluation of the prevalence of non-alcoholic fatty liver disease and steatohepatitis in a large middle-aged US cohort. J Hepatol 2021;75:284–291. doi: 10.1016/j.jhep.2021.02.034

3. Younossi ZM, Henry L. Epidemiology of non-alcoholic fatty liver disease and hepatocellular carcinoma. JHEP Rep 2021;3:100305. doi: 10.1016/j.jhepr.2021.100305

4. Noureddin M, Vipani A, Bresee C, Todo T, Kim IK, Alkhouri N, et al. NASH Leading Cause of Liver Transplant in Women: Updated Analysis of Indications For Liver Transplant and Ethnic and Gender Variances. Am J Gastroenterol 2018;113:1649–1659. doi: 10.1038/s41395-018-0088-6

5. Wegermann K, Hyun J, Diehl AM. Molecular Mechanisms Linking Nonalcoholic Steatohepatitis to Cancer. Clin Liver Dis (Hoboken) 2021;17:6–10. doi: 10.1002/cld.1006

6. Oseini AM, Sanyal AJ. Therapies in non-alcoholic steatohepatitis (NASH). Liver Int 2017;37 Suppl 1:97–103. doi: 10.1111/liv.13302

7. Vuppalanchi R, Noureddin M, Alkhouri N, Sanyal AJ. Therapeutic pipeline in nonalcoholic steatohepatitis. Nat Rev Gastroenterol Hepatol 2021;18:373–392. doi: 10.1038/s41575-020-00408-y

8. Harrison SA, Bashir MR, Guy CD, Zhou R, Moylan CA, Frias JP, et al. Resmetirom (MGL-3196) for the treatment of non-alcoholic steatohepatitis: a multicentre, randomised, double-blind, placebo-controlled, phase 2 trial. Lancet 2019;394:2012–2024. doi: 10.1016/S0140-6736(19)32517-6

9. Harrison SA, Ruane PJ, Freilich B, Neff G, Patil R, Behling C, et al. A randomized, double-blind, placebo-controlled phase IIa trial of efruxifermin for patients with compensated NASH cirrhosis. JHEP Rep 2022;5:100563. doi: 10.1016/j.jhepr.2022.100563

10. Francque SM, Bedossa P, Ratziu V, Anstee QM, Bugianesi E, Sanyal AJ, et al. A Randomized, Controlled Trial of the Pan-PPAR Agonist Lanifibranor in NASH. N Engl J Med 2021;385:1547–1558. doi: 10.1056/NEJMoa2036205

11. Newsome PN, Buchholtz K, Cusi K, Linder M, Okanoue T, Ratziu V, et al. A Placebo-Controlled Trial of Subcutaneous Semaglutide in Nonalcoholic Steatohepatitis. N Engl J Med 2021;384:1113–1124. doi: 10.1056/NEJMoa2028395

12. Loomba R, Sanyal AJ, Kowdley KV, Bhatt DL, Alkhouri N, Frias JP, et al. Randomized, Controlled Trial of the FGF21 Analogue Pegozafermin in NASH. N Engl J Med 2023;389:998–1008. doi: 10.1056/NEJMoa2304286

13. Koutoukidis DA, Mozes FE, Jebb SA, Tomlinson JW, Pavlides M, Saffioti F, et al. A low-energy total diet replacement program demonstrates a favorable safety profile and improves liver disease severity in nonalcoholic steatohepatitis. Obesity (Silver Spring) 2023;31:1767–1778. doi: 10.1002/oby.23793

14. Rinella ME, Neuschwander-Tetri BA, Siddiqui MS, Abdelmalek MF, Caldwell S, Barb D, et al. AASLD Practice Guidance on the clinical assessment and management of nonalcoholic fatty liver disease. Hepatology 2023;77:1797–1835. doi: 10.1097/HEP.0000000000000323

15. Thomaides-Brears HB, Alkhouri N, Allende D, Harisinghani M, Noureddin M, Reau NS, et al. Incidence of Complications from Percutaneous Biopsy in Chronic Liver Disease: A Systematic Review and Meta-Analysis. Dig Dis Sci 2022;67:3366–3394. doi: 10.1007/s10620-021-07089-w

16. Benjamin A, Zubajlo R, Thomenius K, Dhyani M, Kaliannan K, Samir AE, et al. Non-invasive diagnosis of non-alcoholic fatty liver disease (NAFLD) using ultrasound image echogenicity. Annu Int Conf IEEE Eng Med Biol Soc 2017;2017:2920–2923. doi: 10.1109/EMBC.2017.8037468

17. Ratziu V, Charlotte F, Heurtier A, Gombert S, Giral P, Bruckert E, et al. Sampling variability of liver biopsy in nonalcoholic fatty liver disease. Gastroenterology 2005;128:1898–906. doi: 10.1053/j.gastro.2005.03.084

18. Center for Drug Evaluation and Research (CDER) F. Nonalcoholic Steatohepatitis with Compensated Cirrhosis: Developing Drugs for Treatment Guidance for Industry Nonalcoholic Steatohepatitis with Compensated Cirrhosis: Developing Drugs for Treatment Guidance for Industry. Draft Guidance 2019;49:1456–66.

19. Patel J, Bettencourt R, Cui J, Salotti J, Hooker J, Bhatt A, et al. Association of noninvasive quantitative decline in liver fat content on MRI with histologic response in nonalcoholic steatohepatitis. Therap Adv Gastroenterol 2016;9:692–701. doi: 10.1177/1756283X16656735

20. Stine JG, Munaganuru N, Barnard A, Wang JL, Kaulback K, Argo CK, et al. Change in MRI-PDFF and Histologic Response in Patients With Nonalcoholic Steatohepatitis: A Systematic Review and Meta-Analysis. Clin Gastroenterol Hepatol 2021;19:2274–2283.e5. doi: 10.1016/j.cgh.2020.08.061

21. Loomba R, Anstee Q, Harrison S, Sanyal A, Ratziu V, Younossi Z, et al. Obeticholic Acid Improves Hepatic Fibroinflammation as Assessed by Multiparametric Magnetic Resonance Imaging: Interim Results of the REGENERATE Trial - AS075. J Hepatol 2020;73:S19–57. doi: 10.1016/S0168-8278(20)31347-7

22. Tamaki N, Munaganuru N, Jung J, Yonan AQ, Loomba RR, Bettencourt R, et al. Clinical utility of 30% relative decline in MRI-PDFF in predicting fibrosis regression in non-alcoholic fatty liver disease. Gut 2022;71:983–990. doi: 10.1136/gutjnl-2021-324264

23. Imajo K, Tetlow L, Dennis A, Shumbayawonda E, Mouchti S, Kendall TJ, et al. Quantitative multiparametric magnetic resonance imaging can aid non-alcoholic steatohepatitis diagnosis in a Japanese cohort. World J Gastroenterol 2021;27:609–623. doi: 10.3748/wjg.v27.i7.609

24. Permutt Z, Le TA, Peterson MR, Seki E, Brenner DA, Sirlin C, et al. Correlation between liver histology and novel magnetic resonance imaging in adult patients with non-alcoholic fatty liver disease - MRI accurately quantifies hepatic steatosis in NAFLD. Aliment Pharmacol Ther 2012;36:22–9. doi: 10.1111/j.1365-2036.2012.05121.x

25. Wildman-Tobriner B, Middleton MM, Moylan CA, Rossi S, Flores O, Chang ZA, et al. Association Between Magnetic Resonance Imaging-Proton Density Fat Fraction and Liver Histology Features in Patients With Nonalcoholic Fatty Liver Disease or Nonalcoholic Steatohepatitis. Gastroenterology 2018;155:1428–1435.e2. doi: 10.1053/j.gastro.2018.07.018

26. Pickhardt PJ, Hahn L, Muñoz del Rio A, Park SH, Reeder SB, Said A. Natural history of hepatic steatosis: observed outcomes for subsequent liver and cardiovascular complications. AJR Am J Roentgenol 2014;202:752–8. doi: 10.2214/AJR.13.11367

27. Lee SW, Huang DQ, Bettencourt R, Ajmera V, Tincopa M, Noureddin N, et al. Low liver fat in non-alcoholic steatohepatitis-related significant fibrosis and cirrhosis is associated with hepatocellular carcinoma, decompensation and mortality. Aliment Pharmacol Ther 2023. doi: 10.1111/apt.17783

28. Hernando D, van der Heijden RA, Reeder SB. A better understanding of liver T1. Eur Radiol 2023;33:6841–6843. doi: 10.1007/s00330-023-10067-7

29. Banerjee R, Pavlides M, Tunnicliffe EM, Piechnik SK, Sarania N, Philips R, et al. Multiparametric magnetic resonance for the non-invasive diagnosis of liver disease. J Hepatol 2014;60:69–77. doi: 10.1016/j.jhep.2013.09.002

30. Andersson A, Kelly M, Imajo K, Nakajima A, Fallowfield JA, Hirschfield G, et al. Clinical Utility of Magnetic Resonance Imaging Biomarkers for Identifying Nonalcoholic Steatohepatitis Patients at High Risk of Progression: A Multicenter Pooled Data and Meta-Analysis. Clin Gastroenterol Hepatol 2022;20:2451–2461.e3. doi: 10.1016/j.cgh.2021.09.041

31. Pavlides M, Banerjee R, Tunnicliffe EM, Kelly C, Collier J, Wang LM, et al. Multiparametric magnetic resonance imaging for the assessment of non-alcoholic fatty liver disease severity. Liver Int 2017;37:1065–1073. doi: 10.1111/liv.13284

32. Dennis A, Mouchti S, Kelly M, Fallowfield JA, Hirschfield G, Pavlides M, et al. A composite biomarker using multiparametric magnetic resonance imaging and blood analytes accurately identifies patients with non-alcoholic steatohepatitis and significant fibrosis. Sci Rep 2020;10:15308. doi: 10.1038/s41598-020-71995-8

33. Jayaswal ANA, Levick C, Selvaraj EA, Dennis A, Booth JC, Collier J, et al. Prognostic value of multiparametric magnetic resonance imaging, transient elastography and blood-based fibrosis markers in patients with chronic liver disease. Liver Int 2020;40:3071–3082. doi: 10.1111/liv.14625

34. Roca-Fernandez A, Banerjee R, Thomaides-Brears H, Telford A, Sanyal A, Neubauer S, et al. Liver disease is a significant risk factor for cardiovascular outcomes - A UK Biobank study. J Hepatol 2023;79:1085–1095. doi: 10.1016/j.jhep.2023.05.046

35. Harrison SA, Rinella ME, Abdelmalek MF, Trotter JF, Paredes AH, Arnold HL, et al. NGM282 for treatment of non-alcoholic steatohepatitis: a multicentre, randomised, double-blind, placebo-controlled, phase 2 trial. Lancet 2018;391:1174–1185. doi: 10.1016/S0140-6736(18)30474-4

36. Harrison SA, Baum SJ, Gunn NT, Younes ZH, Kohli A, Patil R, et al. Multifactorial effects of AXA1125 and AXA1957 observed on markers of metabolism, inflammation and fibrosis: a 16-week randomized placebo-controlled study in subjects with non-alcoholic fatty liver disease (NAFLD) with and without type 2 diabetes (T2D). Journal of Hepatology 2020;73:S123. doi: 10.1016/S0168-8278(20)30763-7

37. Lawitz EJ, Loomba R, Kowdley KV, Kelly M, Dennis A, Kumar V, et al. Liver-distributed FXR agonist TERN-101 leads to corrected T1 (cT1) response and a population shift to lower cT1 risk categories in NASH phase 2a LIFT study. Journal of Hepatology 2021;74:S1:1875.

38. Tan HC, Shumbayawonda E, Beyer C, Cheng LT, Low A, Lim CH, et al. Multiparametric Magnetic Resonance Imaging and Magnetic Resonance Elastography to Evaluate the Early Effects of Bariatric Surgery on Nonalcoholic Fatty Liver Disease. Int J Biomed Imaging 2023;2023:4228321. doi: 10.1155/2023/4228321

39. Harrison SA, Baum SJ, Gunn NT, Younes ZH, Kohli A, Patil R, et al. Safety, Tolerability, and Biologic Activity of AXA1125 and AXA1957 in Subjects With Nonalcoholic Fatty Liver Disease. Am J Gastroenterol 2021;116:2399–2409. doi: 10.14309/ajg.0000000000001375

40. Loomba R, Abdelmalek MF, Armstrong MJ, Jara M, Kjær MS, Krarup N, et al. Semaglutide 2·4 mg once weekly in patients with non-alcoholic steatohepatitis-related cirrhosis: a randomised, placebo-controlled phase 2 trial. Lancet Gastroenterol Hepatol 2023;8:511–522. doi: 10.1016/S2468-1253(23)00068-7

41. Harrison SA, Rossi SJ, Paredes AH, Trotter JF, Bashir MR, Guy CD, et al. NGM282 Improves Liver Fibrosis and Histology in 12 Weeks in Patients With Nonalcoholic Steatohepatitis. Hepatology 2020;71:1198–1212. doi: 10.1002/hep.30590

42. Ratziu V, Charlton M. Rational combination therapy for NASH: Insights from clinical trials and error. J Hepatol 2023;78:1073–1079. doi: 10.1016/j.jhep.2022.12.025

43. Dennis A, Kelly MD, Fernandes C, Mouchti S, Fallowfield JA, Hirschfield G, et al. Correlations Between MRI Biomarkers PDFF and cT1 With Histopathological Features of Non-Alcoholic Steatohepatitis. Front Endocrinol (Lausanne) 2021;11:575843. doi: 10.3389/fendo.2020.575843

44. Bachtiar V, Kelly MD, Wilman HR, Jacobs J, Newbould R, Kelly CJ, et al. Repeatability and reproducibility of multiparametric magnetic resonance imaging of the liver. PLoS One 2019;14:e0214921. doi: 10.1371/journal.pone.0214921

45. Roskilly A, Hicks A, Taylor EJ, Jones R, Parker R, Rowe IA. Fibrosis progression rate in a systematic review of placebo-treated nonalcoholic steatohepatitis. Liver Int 2021;41:982–995. doi: 10.1111/liv.14749

46. Karlsen TH, Sheron N, Zelber-Sagi S, Carrieri P, Dusheiko G, Bugianesi E, et al. The EASL-Lancet Liver Commission: protecting the next generation of Europeans against liver disease complications and premature mortality. Lancet 2022;399:61–116. doi: 10.1016/S0140-6736(21)01701-3

47. Vali Y, Lee J, Boursier J, Petta S, Wonders K, Tiniakos D, et al. Biomarkers for staging fibrosis and non-alcoholic steatohepatitis in non-alcoholic fatty liver disease (the LITMUS project): a comparative diagnostic accuracy study. Lancet Gastroenterol Hepatol 2023;8:714–725. doi: 10.1016/S2468-1253(23)00017-1

48. Ng CH, Xiao J, Lim WH, Chin YH, Yong JN, Tan DJH, et al. Placebo effect on progression and regression in NASH: Evidence from a meta-analysis. Hepatology 2022;75(6):1647–1661. doi: 10.1002/hep.32315

49. Noureddin M, Ntanios F, Malhotra D, Hoover K, Emir B, McLeod E, et al. Predicting NAFLD prevalence in the United States using National Health and Nutrition Examination Survey 2017-2018 transient elastography data and application of machine learning. Hepatol Commun 2022;6:1537–1548. doi: 10.1002/hep4.1935

50. Beyer C, Andersson A, Shumbayawonda E, Dennis A, Corey K. Assessing the repeatability of LiverMultiScan metric cT1 in patients with nonalcoholic steatohepatitis [abstract]. In: Paris NASH Meeting; 2023 Sep 7-8; Paris, France

