## Supplementary tables for "Decreases in liver cT1 accurately reflect histological improvement induced by therapies in MASH: a multi-centre pooled cohort analysis" (from another version of this preprint: version 1, 2024-02-29)

**Supplementary Material**

**Supplementary Table 1: Descriptive statistics table for MRI and histology metrics at baseline and follow-up timepoint for each of the individual included studies.** Normally distributed metrics are presented as mean (standard deviation), and non-normally distributed metrics are presented as median [IQR].

|  | **Study 1**  **(n = 18)** | | | **Study 2**  **(n = 28)** | | | **Study 3**  **(n = 104)** | | |
| --- | --- | --- | --- | --- | --- | --- | --- | --- | --- |
|  | **Baseline** | **Follow-up** | **P-value** | **Baseline** | **Follow-up** | **P-value** | **Baseline** | **Follow-up** | **P-value** |
| Imaging endpoints | | | | | | | | | |
| **cT1 (ms)** | 904 [47.8] | 814 [55.8] | **<0.001** | 903 [122.8] | 780 [101.8] | **<0.001** | 934.5 [139] | 892 [154.5] | 0.053 |
| **PDFF (%)** | 16.9 [7.3] | 4.7 [4.1] | **<0.001** | 19 [6.7] | 5.7 [3.9] | **<0.001** | 18.9 [7.5] | 16.3 [9.1] | **<0.001** |
| **Components of histology scoring** | | | | | | | | | |
| **Steatosis** | 1.6 (0.8) | 0.5 (0.5) | **<0.001** | 2.2 (0.7) | 0.9 (1) | **<0.001** | 1.9 (0.7) | 1.6 (0.9) | **0.018** |
| **Inflammation** | 2.2 (0.6) | 1.8 (0.5) | 0.056 | 1.9 (0.3) | 1 (0.6) | **<0.001** | 1.1 (0.5) | 1 (0.6) | 0.61 |
| **Ballooning** | 1.8 (0.5) | 1.2 (0.8) | **0.0087** | 1.4 (0.5) | 0.5 (0.6) | **<0.001** | 1.4 (0.6) | 1.2 (0.6) | **0.018** |
| **NAS** | 5.6 (1.4) | 3.6 (1.3) | **<0.001** | 5.5 (0.8) | 2.5 (1.6) | **<0.001** | 4.4 (1.1) | 3.9 (1.3) | **0.0069** |
| **Fibrosis** | 2.5 (0.9) | 2.1 (0.9) | 0.13 | 2 (0.9) | 1.4 (1.3) | **0.042** | 2.4 (0.8) | 2.6 (1) | 0.15 |

**Supplementary Table 2: Score distribution table for all histology metrics at baseline and follow-up timepoint for the total cohort, and each of the individual included studies.** Each score is reported as total number, and percentage of all patients per group.

|  | **Total Cohort**  **(n = 150)** | | **Study 1**  **(n = 18)** | | **Study 2**  **(n = 28)** | | **Study 3**  **(n = 104)** | |
| --- | --- | --- | --- | --- | --- | --- | --- | --- |
|  | **Baseline** | **Follow-up** | **Baseline** | **Follow-up** | **Baseline** | **Follow-up** | **Baseline** | **Follow-up** |
| **Steatosis**  Score (n, %) | 0 (0, 0%);  1 (48, 32%);  2 (66, 44%);  3 (36, 24%); | 0 (29, 19.3%);  1 (59, 39.3%);  2 (41, 27.3%);  3 (21, 14%); | 0 (0, 0%);  1 (11, 61.1%);  2 (4, 22.2%);  3 (3, 16.7%); | 0 (9, 50%);  1 (9, 50%);  2 (0, 0%);  3 (0, 0%); | 0 (0, 0%);  1 (4, 14.3%);  2 (15, 53.6%);  3 (9, 32.1%); | 0 (11, 39.3%);  1 (11, 39.3%);  2 (3, 10.7%);  3 (3, 10.7%); | 0 (0, 0%);  1 (33, 31.7%);  2 (47, 45.2%);  3 (24, 23.1%); | 0 (9, 8.7%);  1 (39, 37.5%);  2 (38, 36.5%);  3 (18, 17.3%); |
| **Inflammation**  Score (n, %) | 0 (10, 6.7%);  1 (81, 54%);  2 (52, 34.7%);  3 (7, 4.7%); | 0 (18, 12%);  1 (96, 64%);  2 (34, 22.7%);  3 (2, 1.3%); | 0 (0, 0%);  1 (2, 11.1%);  2 (10, 55.6%);  3 (6, 33.3%); | 0 (0, 0%);  1 (4, 22.2%);  2 (13, 72.2%);  3 (1, 5.6%); | 0 (0, 0%);  1 (2, 7.1%);  2 (26, 92.9%);  3 (0, 0%); | 0 (4, 14.3%);  1 (19, 67.9%);  2 (5, 17.9%);  3 (0, 0%); | 0 (10, 9.6%);  1 (77, 74%);  2 (16, 15.4%);  3 (1, 1%); | 0 (14, 13.5%);  1 (73, 70.2%);  2 (16, 15.4%);  3 (1, 1%); |
| **Ballooning**  Score (n, %) | 0 (4, 2.7%);  1 (73, 48.7%);  2 (73, 48.7%); | 0 (30, 20%);  1 (78, 52%);  2 (42, 28%); | 0 (1, 5.6%);  1 (2, 11.1%);  2 (15, 83.3%); | 0 (4, 22.2%);  1 (7, 38.9%);  2 (7, 38.9%); | 0 (0, 0%);  1 (17, 60.7%);  2 (11, 39.3%); | 0 (16, 57.1%);  1 (10, 35.7%);  2 (2, 7.1%); | 0 (3, 2.9%);  1 (54, 51.9%);  2 (47, 45.2%); | 0 (10, 9.6%);  1 (61, 58.7%);  2 (33, 31.7%); |
| **NAS**  Score (n, %) | 0 (0, 0%);  1 (0, 0%);  2 (4, 2.7%);  3 (20, 13.3%);  4 (36, 24%);  5 (52, 34.7%);  6 (28, 18.7%);  7 (7, 4.7%);  8 (3, 2%); | 0 (4, 2.7%);  1 (9, 6%);  2 (22, 14.7%);  3 (34, 22.7%);  4 (38, 25.3%);  5 (28, 18.7%);  6 (15, 10%);  7 (0, 0%);  8 (0, 0%); | 0 (0, 0%);  1 (0, 0%);  2 (0, 0%);  3 (2, 11.1%);  4 (0, 0%);  5 (8, 44.4%);  6 (4, 22.2%);  7 (2, 11.1%);  8 (2, 11.1%); | 0 (0, 0%);  1 (1, 5.6%);  2 (4, 22.2%);  3 (1, 5.6%);  4 (9, 50%);  5 (2, 11.1%);  6 (1, 5.6%);  7 (0, 0%);  8 (0, 0%); | 0 (0, 0%);  1 (0, 0%);  2 (0, 0%);  3 (0, 0%);  4 (3, 10.7%);  5 (11, 39.3%);  6 (11, 39.3%);  7 (3, 10.7%);  8 (0, 0%); | 0 (3, 10.7%);  1 (4, 14.3%);  2 (10, 35.7%);  3 (5, 17.9%);  4 (2, 7.1%);  5 (2, 7.1%);  6 (2, 7.1%);  7 (0, 0%);  8 (0, 0%); | 0 (0, 0%);  1 (0, 0%);  2 (4, 3.8%);  3 (18, 17.3%);  4 (33, 31.7%);  5 (33, 31.7%);  6 (13, 12.5%);  7 (2, 1.9%);  8 (1, 1%); | 0 (1, 1%);  1 (4, 3.8%);  2 (8, 7.7%);  3 (28, 26.9%);  4 (27, 26%);  5 (24, 23.1%);  6 (12, 11.5%);  7 (0, 0%);  8 (0, 0%); |
| **Fibrosis**  Stage (n, %) | 0 (0, 0%);  1 (28, 18.7%);  2 (50, 33.3%);  3 (64, 42.7%);  4 (8, 5.3%); | 0 (12, 8%);  1 (23, 15.3%);  2 (43, 28.7%);  3 (54, 36%);  4 (18, 12%); | 0 (0, 0%);  1 (3, 16.7%);  2 (4, 22.2%);  3 (10, 55.6%);  4 (1, 5.6%); | 0 (0, 0%);  1 (6, 33.3%);  2 (6, 33.3%);  3 (5, 27.8%);  4 (1, 5.6%); | 0 (0, 0%);  1 (10, 35.7%);  2 (8, 28.6%);  3 (10, 35.7%);  4 (0, 0%); | 0 (8, 28.6%);  1 (9, 32.1%);  2 (4, 14.3%);  3 (5, 17.9%);  4 (2, 7.1%); | 0 (0, 0%);  1 (15, 14.4%);  2 (38, 36.5%);  3 (44, 42.3%);  4 (7, 6.7%); | 0 (4, 3.8%);  1 (8, 7.7%);  2 (33, 31.7%);  3 (44, 42.3%);  4 (15, 14.4%); |
